# Epigenomics and Transcriptomics of Systemic Sclerosis CD4+ T cells reveal Long Range Dysregulation of Key Inflammatory Pathways mediated by disease-associated Susceptibility Loci

**DOI:** 10.1101/2020.03.14.20036061

**Authors:** Tianlu Li, Lourdes Ortiz, Eduardo Andrés-León, Laura Ciudad, Biola M. Javierre, Elena López-Isac, Alfredo Guillén-Del-Castillo, Carmen Pilar Simeón-Aznar, Esteban Ballestar, Javier Martin

## Abstract

System sclerosis (SSc) is a genetically complex autoimmune disease mediated by the interplay between genetic and epigenetic factors in a multitude of immune cells, with CD4+ T lymphocytes as one of the principle drivers of pathogenesis. In this study, we obtained DNA methylation and expression profiles of CD4+ T cells from 48 SSc patients and 16 healthy controls. Consequently, we identified 9112 and 3929 differentially methylated CpGs positions (DMPs) and differentially expressed genes (DEGs) respectively. These DMPs and DEGs are enriched in functional categories related to inflammation and T cell biology. Furthermore, correlation analysis identified 17,500 possible DMP-DEG interaction pairs within a window of 5 Mb, and utilizing promoter capture Hi-C data, we confirmed that 212 CD4+ T cel specific pairs of DMP-DEG physically interact involving CTCF. Finally, utilizing SSc GWAS data, we identified four important SSc-associated susceptibility loci, *TNIP1* (rs3792783), *GSDMB* (rs9303277), *IL12RB1* (rs2305743) and *CSK* (rs1378942), that physically interact with DMP-DEG pairs cg17239269-*ANXA6*, cg19458020-*CCR7*, cg10808810-*JUND* and cg11062629-*ULK3* respectively. Overall, our study reveals a solid link between genetic, epigenetic and transcriptional deregulation in CD4+ T cells of SSc patients, providing a novel integrated view of SSc pathogenic determinants.

## INTRODUCTION

Systemic sclerosis (SSc) is a chronic, progressive autoimmune disease of unknown etiology that is primarily characterised by extensive microvascular damage, deregulation of immune cells and systemic fibrosis of skin and various organs. The estimated overall 10-year survival rate is 66 %, which significantly decreases upon organ involvement ^1^. SSc patients can be broadly categorized into two groups based on the extent of skin involvement: those with restricted involvement affecting the limbs distal to the elbows or knees with o without face and neck involvement are classified as limited cutaneous SSc or those with proximal involvement affecting above the elbows and knees are classified as diffuse cutaneous SSc ^2^. The pathogenesis of SSc is not well-understood, in which disease onset and development appear to be multistep and multifactorial processes involving both genetic and environmental factors ^3,4^.

Like most autoimmune diseases, genetics studies have revealed that SSc heritability is complex with the HLA loci playing an important contribution to disease risk ^5^. Recent genome-wide association (GWAS) and Immunochip array SNP studies revealed the presence of numerous non-HLA loci that associate with disease onset, including genes of type I interferon pathway, interleukin-12 pathway, TNF pathway as well as B and T cell specific genes ^6–12^. Furthermore, recent chromatin interaction analyses have linked these genetic variants to potential target genes implicated in SSc disease progression ^13^. However, genetic variants do not account for all of the genetic burden of SSc, and other factors, such as epigenetic dysregulation, play an indispensable role in disease pathogenesis ^3,14^.

It is well-established that CD4+ T cells play a pivotal role in the pathogenesis of several autoimmune diseases. Abnormalities in the proportions of CD4+ T lymphocyte subpopulations were detected more than two decades ago in SSc patients ^15,16^, and since then, several studies proposed mechanistic alterations in these lymphocyte populations that may contribute to disease manifestations. Firstly, increased production of several CD4+ T cell-mediated cytokines, including IL-27, IL-6, TGF-β and IL-17A, were detected in the serum of SSc patients, which may directly drive vascular dysregulation and fibrosis ^17,18^. Secondly, CD4+ T cells isolated from SSc patients were observed to be functionally impaired, as stimulation resulted in deregulated polarization towards Th17 expansion, as well as inherent diminished immune capacity of circulating Treg cells ^19,20^. Finally, aberrant interactions with other cell types, including mesenchymal stromal cells and fibroblasts, have been observed ^21,22^. The exact mechanism that drive CD4+ T cell deregulation in SSc is currently unknown, however, there is strong evidence that alterations in DNA methylation may be a primary culprit. DNA methylation plays a critical role in T cell polarization and activation, in which naïve T cells undergo reprograming of its methylome to increase accessibility of selective loci upon differentiation into different T-helper lymphocyte subpopulations (reviewed in ^23,24^). Gene expression of DNA methylation-related proteins were observed to be deregulated in SSc ^25^. Several studies have observed aberrant methylation of promoters of such genes as *CD40L* ^26^, *TNFSF7* ^27^, *FOXP3* ^28^, and IFN-associated genes ^29^, which may result in their aberrant expression.

In this study, we describe the direct relationship between DNA methylation and gene expression in SSc CD4+ T cells, and how aberrant DNA methylation potentially deregulates the expression of several important inflammatory genes through long distance physical interactions, that involve CTCF. Furthermore, these alterations may be a direct result of the presence of genetic variants in nearby loci.

## RESULTS

### DNA methylation deregulation in SSc CD4 T cells

To gain insights into functional and molecular alterations of T lymphocytes in the context of SSc pathogenesis, we first isolated CD4+ T lymphocytes from the PBMCs of 48 SSc patients and 16 age- and sex-matched healthy donors (HD) by CD4+ positive cell sorting (Figure 1A and B and Supplementary Table 1). To interrogate DNA methylation, we utilized bead arrays (see Materials and Methods). Subsequently, 9112 CpGs were found to be differentially methylated in SSc CD4+ T cells compared to HD (FDR < 0.05 and p-value < 0.01), in which 7837 and 1275 CpGs were hyper- and hypomethylated respectively (Figure 1C and Supplementary Table 2). Analysis of <u>D</u>ifferentially <u>M</u>ethylated CpG <u>P</u>ositions (DMPs) was visualized by t-Distributed Stochastic Neighbour Embedding (t-SNE) analysis (Figure 1D), and HD and SSc samples can be observed to separate along the t-SNE2 axis. DMPs were predominantly situated in open sea and intergenic regions (Supplementary Figure 1A). Gene ontology analysis revealed relevant categories for both hyper- and hypomethylated DMPs (Figure 1E). Hypermethylated DMPs were enriched in inflammatory pathways, including IL-23 and IL-18 receptor activity and TLR3 signaling pathway, as well as pathways involved in T cell biology, such as memory and Th17 T cell differentiation (Figure 1E, upper panel). On the other hand, hypomethylated DMPs were predominantly enriched in genes encoding the MHC protein complex (Figure 1E, lower panel and Supplementary Figure 1B). Other relevant categories, including proliferation, Th1-type immune response and IL-10 production, were also enriched in hypomethylated DMPs. Motif enrichment analyses revealed zinc finger transcription factor CTCF to be common to both hyper- and hypomethylated DMPs (Figure 1F). This observation is especially interesting given the importance of CTCF in mediating enhancer-gene interactions ^30^, hence aberrant DNA methylation of CTCF-binding enhancer regions may affect the expression of interacting genes. Furthermore, other motifs of TF complexes important to T cell biology, such as the BAFT-JUN-AP1 complex, shown to play a role in CD4+ T cell differentiation ^31^, and TFs of the ETS family, whose deletion in CD4+ T cells result in autoimmunity in mice ^32^, were enriched in the hypomethylated cluster (Figure 1F). Subsequent analyses revealed that hypermethylated DMPs showed a particular histone mark signature that was enriched in H3K9me3, H3K27me3 and H3K4me1 (Figure 1G), which suggest the enrichment of poised enhancers, as was previously defined by Zentner *et al*. ^33^. Conversely, hypomethylated DMPs were significantly enriched in H3K4me1 only, which is a hallmark of primed enhancers (Figure 1G) ^34^. Finally, among the DNA methylation alterations detected in SSc patients, we also observed 852 CpGs that changed their variance compared to healthy controls, and these are termed differentially variable positions (DVPs; Figure 1H). Many of these CpGs were also differentially methylated, however, 453 CpGs were exclusively DVPs, and majority of them experienced an increase in variance in SSc patients (Figure 1H). Increases in variance may be a consequence of differences in pathological evolution of the disease in the patient population. Overall, deregulation in DNA methylation of genes important to immune response and T cell biology were detected in peripheral blood-isolated CD4+ T cells.

**Figure 1.**
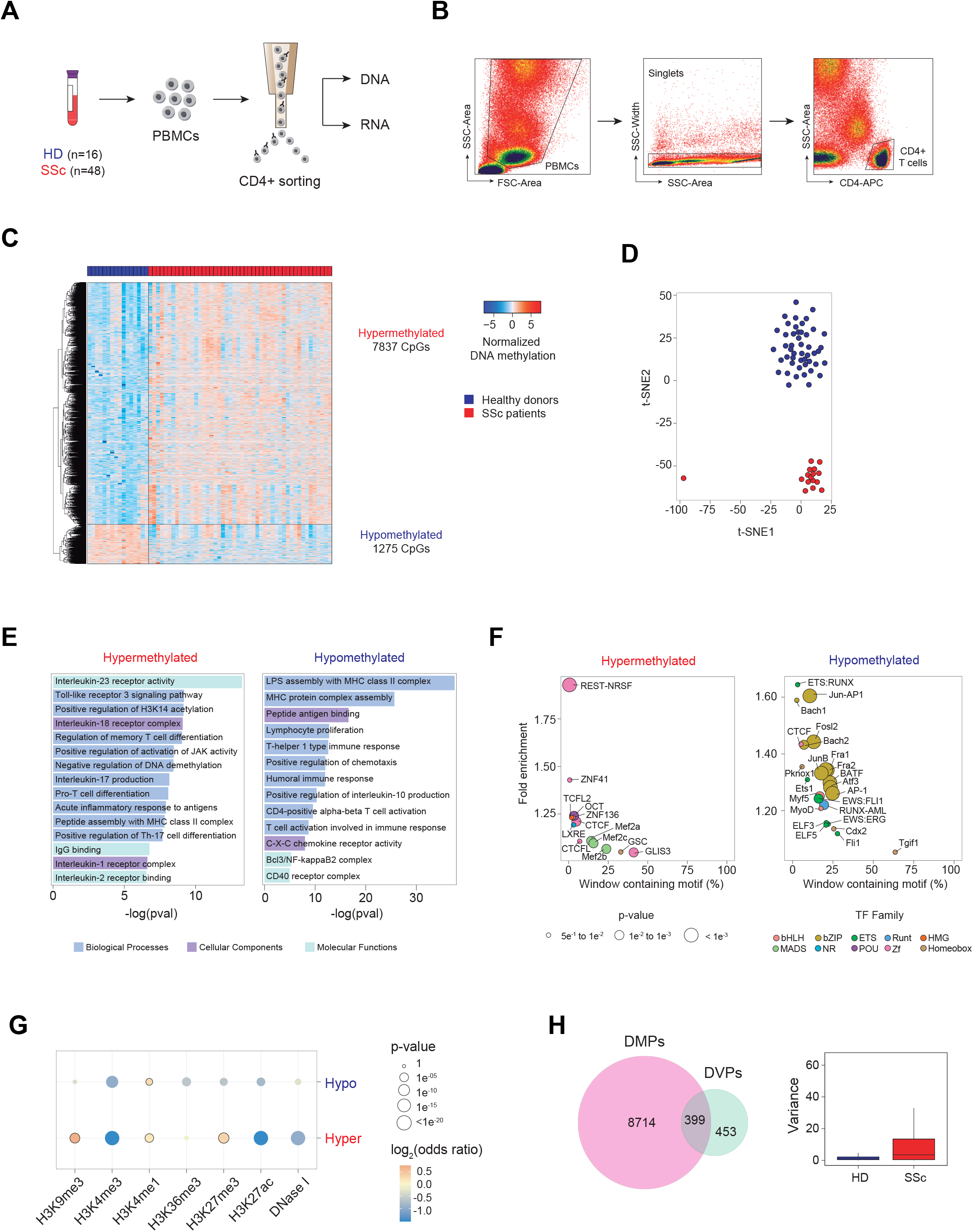
Aberrant DNA methylation in inflammatory loci of SSc CD4+ T cells. (A) Scheme depicting workflow including PBMC isolation, CD4+ T cell positive sorting and DNA and RNA extraction. (B) Gating strategy to eliminate cell debris, doublets and the isolation of CD4-APC+ T lymphocytes by side scatter. (C) Heatmap of differential methylated CpGs (DMPs), in which 7,837 CpGs were hypermethylated (log2FC > 0, FDR < 0.05) and 1,275 CpGs were hypomethylated (log2FC < 0, FDR < 0.05) in SSc patients (n = 48) compared to healthy control (n = 16). (D) t-SNE clustering of SSc and HD samples based on all identified DMPs. (E) Gene ontology of hyper-and hypomethylated DMPs, analyzed utilizing the GREAT online tool (http://great.stanford.edu/), in which CpGs annotated in the EPIC array were used as background. (F) HOMER motif enrichment of hyper-and hypomethylated CpGs, utilizing CpGs annotated in the EPIC array as background. A window of ±250 bp centering around each DMP was applied. (G) Enrichment of DNase I hypersensitivity, H3K9me3, H3K4me3, H3K4me1, H3K36me3, H3K27me3 and H3K27ac ChIP-seq data, obtained from the BLUEPRINT portal, in hyper- and hypomethylated DMPs. Highlighted circles represent statistically significant comparisons (p-value < 0.01 and odds ratio > 1) compared to background. (H) Overlap between DMPs and differentially variable CpGs (DVPs), identified utilizing the iEVORA algorithm (left), and graphical representation of variance of identified DVPs in HD and SSc.

SSc patients were then stratified as diffuse (dcSSc), limited (lcSSc) or sine scleroderma (ssSSc), with the latter having no skin fibrosis. We analyzed aberrant DNA methylation of SSc subgroups and observed that many of the alterations (35 %) were shared between at least two of the three subgroups (Supplementary Figure 1C). Furthermore, subgroup-specific DMPs also displayed some degree of aberrancy in other SSc subgroups that did not reach statistical significance (Supplementary Figure 1D), suggesting that the majority of alterations are shared amongst groups. Altogether, these results suggest that aberrant DNA methylation of CD4+ T cells is a common hallmark of all subgroups of SSc.

### Differentially methylated regions enriches in NF-kB signalling pathway

Although the identification of single DMPs can hint at possible alterations in the genomic landscape, differentially methylated regions (DMRs) may give functional relevance to DNA methylation, as DMRs have been described to correlate well with transcription, chromatin features and phenotypic outcomes ^35,36^. Accordingly, we identified 1082 significant DMRs, in which 212 and 870 DMRs were hypo- and hypermethylated respectively. More than 25 % of DMRs were found to be situated in promoters of the nearest gene, and the same proportion of DMRs were found more than 50 kb from TSS (Supplementary Figure 2A). Many of the identified DMRs were mapped to such relevant genes as *COLEC11, GSTM1, HLA-C* and *IL15RA* (Figure 2A). Gene ontology analyses revealed that hypermethylated DMRs were significantly enriched in various categories relevant to immune functions, IL-10 secretion, Th2 cytokine production and differentiation, NLRP3 inflammasome complex and negative regulation of NF-κB signalling (Figure 2B). On the other hand, hypomethylated DMRs were enriched in peptide antigen binding, MHC protein complex, IL-1 binding and chemotaxis (Figure 2B). Furthermore, both hyper- and hypomethylated DMRs overlapped significantly with H3K4me1 histone mark and DNase I hypersentivity regions, indicating the presence of enhancers (Figure 2C). Additionally, hypermethylated DMRs were enriched in H3K27me3, which, together with H3K4me1, is a key mark for poised enhancers. Given the importance of CTCF in mediating long-range enhancer-DNA interactions and the enrichment of its motif in both hyper- and hypomethylated DMPs, we performed overlap between DMRs and CTCF and CTCFL ChIP-seq peaks obtained from ENCODE. We observed that hyperDMRs significantly overlapped with both CTCF and CTCFL binding regions (Figure 2D). Moreover, significant overlap was also detected between both hyper- and hypoDMRs and binding sites of RUNX1, MYC and RELA, all of which have important roles in T cell biology, suggesting that aberrant DNA methylation in SSc may alter their binding to these regions. Finally, hypoDMRs were selectively enriched in binding sites of NFKB2 and JUN (Figure 2D).

**Figure 2.**
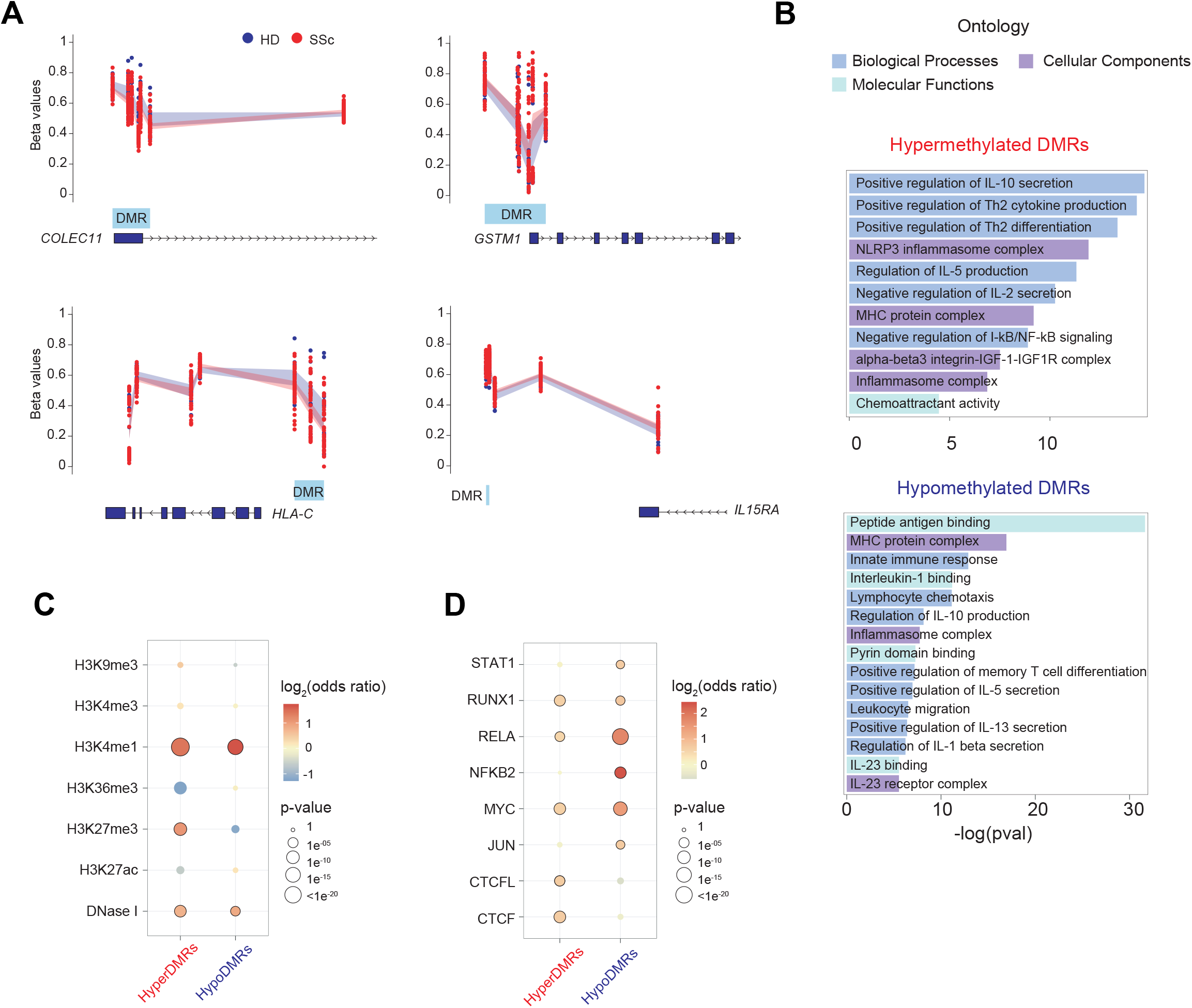
Differentially methylated regions (DMRs) enrich in inflammatory loci and pro-inflammatory transcription factors. (A) Graphical representation of beta values of identified DMRs, utilizing *bumphunter* tool, mapped to relevant genes, including *COLEC11, GSTM1, HLA-C* and *IL15RA*. Identified DMRs are highlighted in blue. (B) GO analysis of hyper- and hypomethylated DMRs utilizing the GREAT online tool. DMRs were mapped to the nearest gene. (C) Enrichment of DNase I hypersensitivity, H3K9me3, H3K4me3, H3K4me1, H3K36me3, H3K27me3 and H3K27ac ChIP-seq data, obtained from the BLUEPRINT portal, in hyper- and hypomethylated DMRs. (D) Enrichment of TF ChIP-seq peaks in hyper- and hypomethylated DMRs. ChIP-seq data were downloaded from ReMap database, in which STAT1, RUNX1, NFKB2, MYC, JUN, CTCFL and CTCF were downloaded as consolidated ChIP-seq peaks of all available datasets, whereas RELA ChIP-seq peaks were obtained from CD4+ T cells. (C-D). Background of the same length as DMRs were generated from EPIC array, in which 1000 resampling was applied. P-values and odds ratios are averages of 1000 permutations. Highlighted circles represent statistically significant comparisons (p-value < 0.01 and odds ratio > 1) compared to background.

### Identification of aberrant gene expression in CD4 T cells of SSc patients

To characterize gene expression aberrancies in CD4+ T cells that drive disease pathogenesis of SSc, we performed genome-wide RNA expression analysis, and found that a total 3929 genes displayed differential expression (Differentially Expressed Genes: DEGs) between HD and SSc, of which 1949 and 1980 were down- and upregulated respectively (Figure 3A, 3B and Supplementary Table 3). To identify the specific pathways that were altered, we performed DAVID gene ontology analysis and observed that downregulated genes were enriched in such signalling pathways as T cell receptor, IL-2 production, Fc-γ receptor and interferon-gamma, as well as genes associated with systemic lupus erythematosus and viral carcinogenesis (Figure 3C). Conversely, genes encoding proteins that propagate signalling pathways such as TNF, NF-κB, Fc-ε receptor, Wnt and TLR-9 were upregulated (Figure 3C). TF enrichment analysis revealed that upregulated DEGs were driven by such TFs as IRF2, NFKB1, FOXP1 and SOX10, and downregulated DEGs were regulated by TFs including EGR1, FOXA1, GATA2/3 and CTCF (Figure 3D). Furthermore, the expression of several transcripts of the HLA cluster were also differentially expressed in SSc patients (Figure 3E), which was in accordance with observed aberrant DNA methylation in these genomic regions. Interestingly, genes encoding several transcription factors whose motifs were enriched in aberrant DMPs were also found to be altered, and these genes include *JUN, FLI1, RUNX1* and *CTCF* (Figure 3E). The expression of other relevant TFs, such as *NFKB2, EGR1* and *RELB* were also dysregulated (Figure 3E).

**Figure 3.**
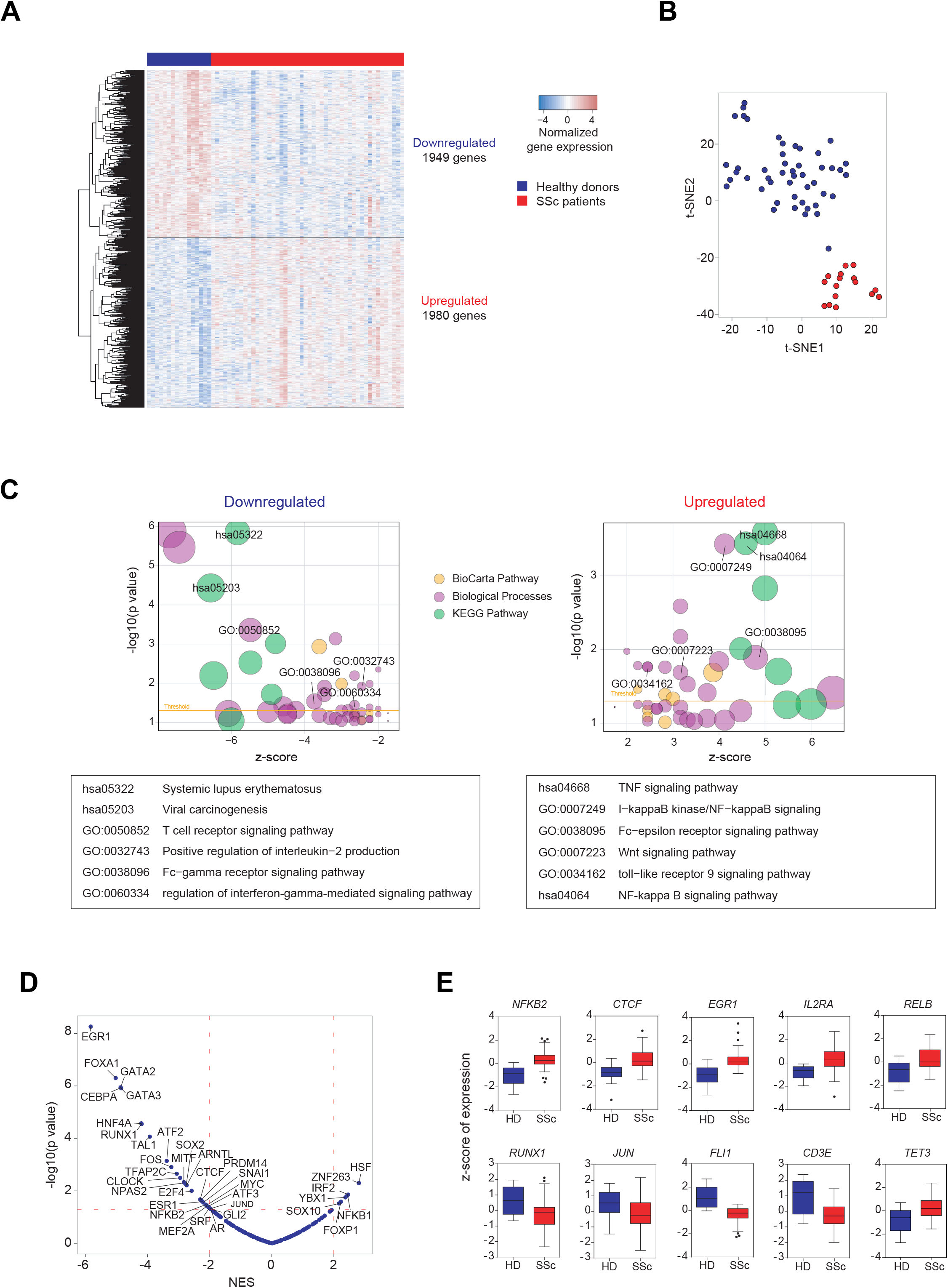
Aberrant gene expression of SSc CD4+ T cells include inflammatory genes and relevant transcription factors. (A) Differentially expression genes (DEGs) comparing RNA expression microarray data of CD4+ T cells isolated from SSc with HD, in which 1,949 genes were downregulated (log_2_FC < 0, FDR < 0.05) and 1,980 genes were upregulated (log_2_FC > 0, FDR < 0.05). (B) t-SNE clustering of identified DEGs in HD and SSc CD4 T cell samples. (C) GO analysis of down- and upregulated DEGs performed utilizing the DAVID online tool (https://david.ncifcrf.gov/). Bubble size refer to size of GO term and threshold represent the cut-off for statistical significance (p-value < 0.05). Relevant GO terms are summarized in tables before the bubble plots. (D) TF enrichment analysis, utilizing the DoRaThea tool, of all interrogated genes ordered by Normalized Enrichment Score (NES) of SSc compared to HD. Dotted red lines represent the cut-offs for statistical significance (NES < -2 & p-value < 0.01, NES > 2 & p-value < 0.01). (E) Graphical representations of z-scores of genes relevant to T cell biology, including *CD3E* and *IL2RA*, transcription factors *NFKB2, CTCF, EGR1, RELB, RUNX1, JUN* and *FLI1*, and DNA methylcytosine dioxygenase *TET3*.

### DNA methylation changes establish short and long distance relationships with gene expression in SSc

We then investigated the relationship between DNA methylation and gene expression deregulation in SSc. TF binding can be both negatively or positively influenced by DNA methylation ^37,38^. DNA methylation of gene TSS associate negatively with gene expression ^39^, whereas methylation of CpGs located within gene body can also associate with active gene expression ^40^. To investigate the potential causal relationship between DNA methylation alterations and gene expression in SSc, we searched for possible meQTLs (methylation-expression quantitative trait loci). Subsequently, we found 45 differentially methylated CpGs (DMPs) that interacted with differentially expressed genes (DEGs), in which the CpG was located near/in the promoter or TSS of the interacting gene (5 kb upstream and 1kb downstream of TSS), of which, 33 (73.3 %) were negative interactions (Supplementary Table 4; Figure 4A). GO analysis of interacting DEGs include pathways involved in nucleotide-excision repair, nucleosome assembly and positive regulation of interferon-beta production (Figure 4B). Relevant genes include *ADAM20*, a member of the ADAM family of metalloproteases which are involved in T cell responses ^41,42^, *CD274*, which, upon binding to its receptor, mediates changes in T cell metabolism ^43^, and *IRF1*, a key driver of Th1 cell differentiation ^44^ (Figure 4C).

**Figure 4.**
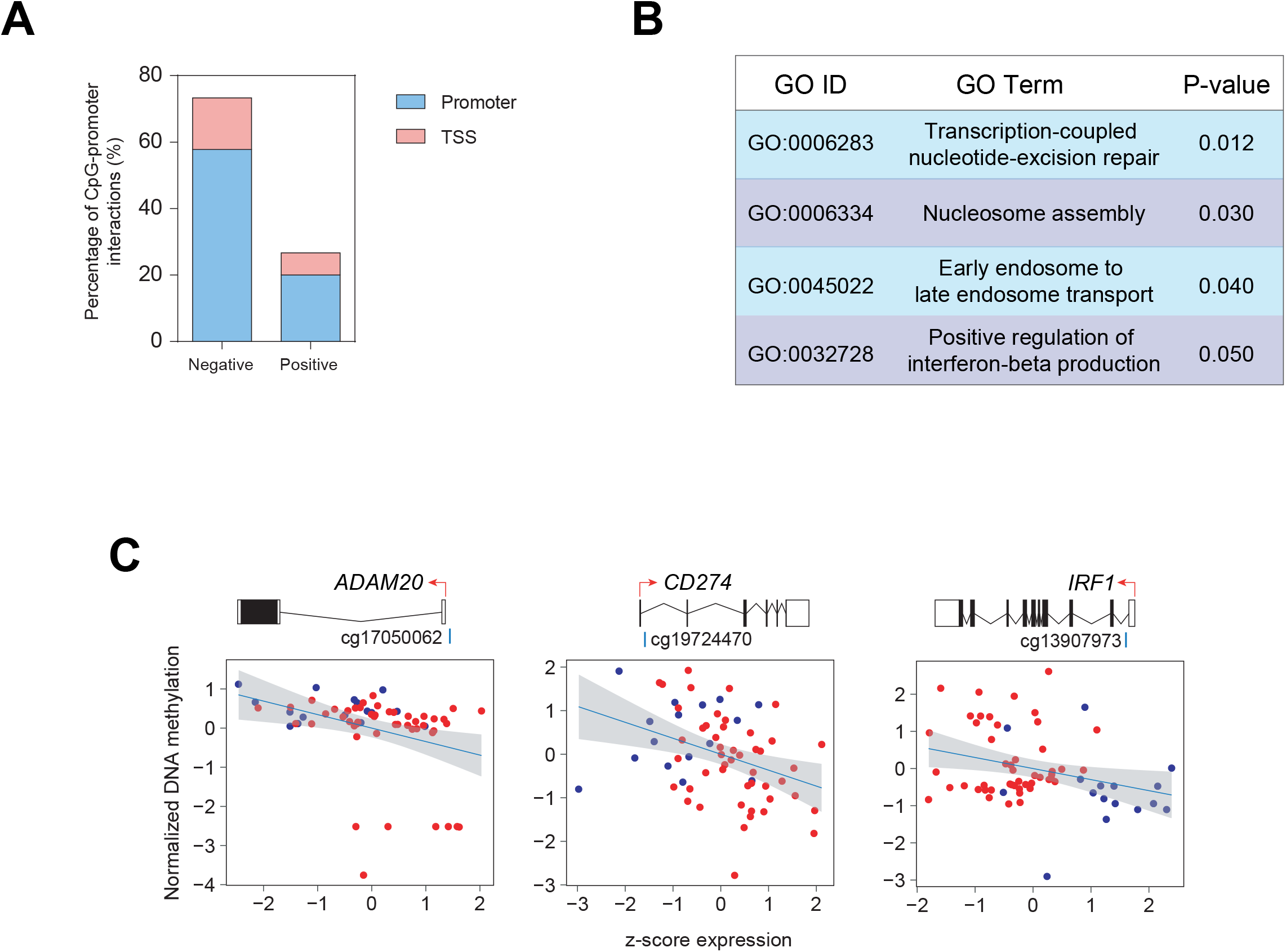
Correlation of DNA methylation and gene expression of DMPs situated in gene promoters/TSS. (A) Percentage of detected short-range interactions that present either negative or position correlation between DNA methylation and gene expression, in which the DMP is situated in promoter or TSS of interacting DEG. (B) GO analysis utilizing the DAVID online tool of the DEGs with short-range interacting DMPs. (C) Graphical representation of correlation of DNA methylation and gene expression of DMP-DEG pairs. Red and blue dots represent SSc and HD individuals respectively.

Given that SSc-associated differentially methylated CpGs and regions (DMPs and DMRs) were enriched in H3K4me1, a key histone mark characterizing enhancers, it is possible that aberrant expression of genes in SSc patient CD4+ T cells may be mediated by CpGs situated in distal elements. To interrogate long distance interactions, an extended window of 5 Mb between CpG and gene was used to detect meQTLs followed by extensive multi-step filtering processes based on the causal inference test ^45,46^ (Figure 5A). Firstly, correlations between all annotated CpG positions (M) and genes (E) were searched within a window of 5 Mb, in which a total of 99,525 significant (Pearson p-value < 0.01) CpG-gene interactions were discovered. Secondly, CpG-gene pairs were then filtered by significant association with SSc (Y) to yield 17,500 DMP-DEG pairs (5,841 unique DMPs and 3,237 unique genes), approximately half (9,114 DMP-DEG pairs) of which were negative correlations. Thirdly, utilizing previously generated promoter capture Hi-C (PCHi-C) data of CD4+ T cells ^47^, we overlapped promoter-non-promoter interactions with SSc-associated DMP-DEG pairs. Consequently, our analysis yielded 182, 170 and 162 DMP-DEG interactions utilizing datasets from naïve, non-activated and activated CD4+ T cells, respectively. Following consolidation of the three datasets, we detected a total of 212 unique DMP-DEG interactions (Supplementary Table 5), in which 128 interactions were shared between all three datasets (Figure 5B). Furthermore, these interactions appeared to be specific to CD4+ T cells, as comparison with PCHi-C data obtained from erythroblasts displayed little overlap (Figure 5A). Furthermore, of the 212 confirmed DMP-DEG interactions, more than half displayed a negative correlation between gene expression and DNA methylation (Figure 5C and D). Among these confirmed DMP-DEG interactions, aberrantly expressed genes relevant to T cell biology, such as *ANXA6, CCR7, CD274, CD4, CD48, IRAK2, JUND* and *NFKB2*, were shown to directly interact with CpGs that displayed differential DNA methylation in SSc patients (Figure 5D-F).

**Figure 5.**
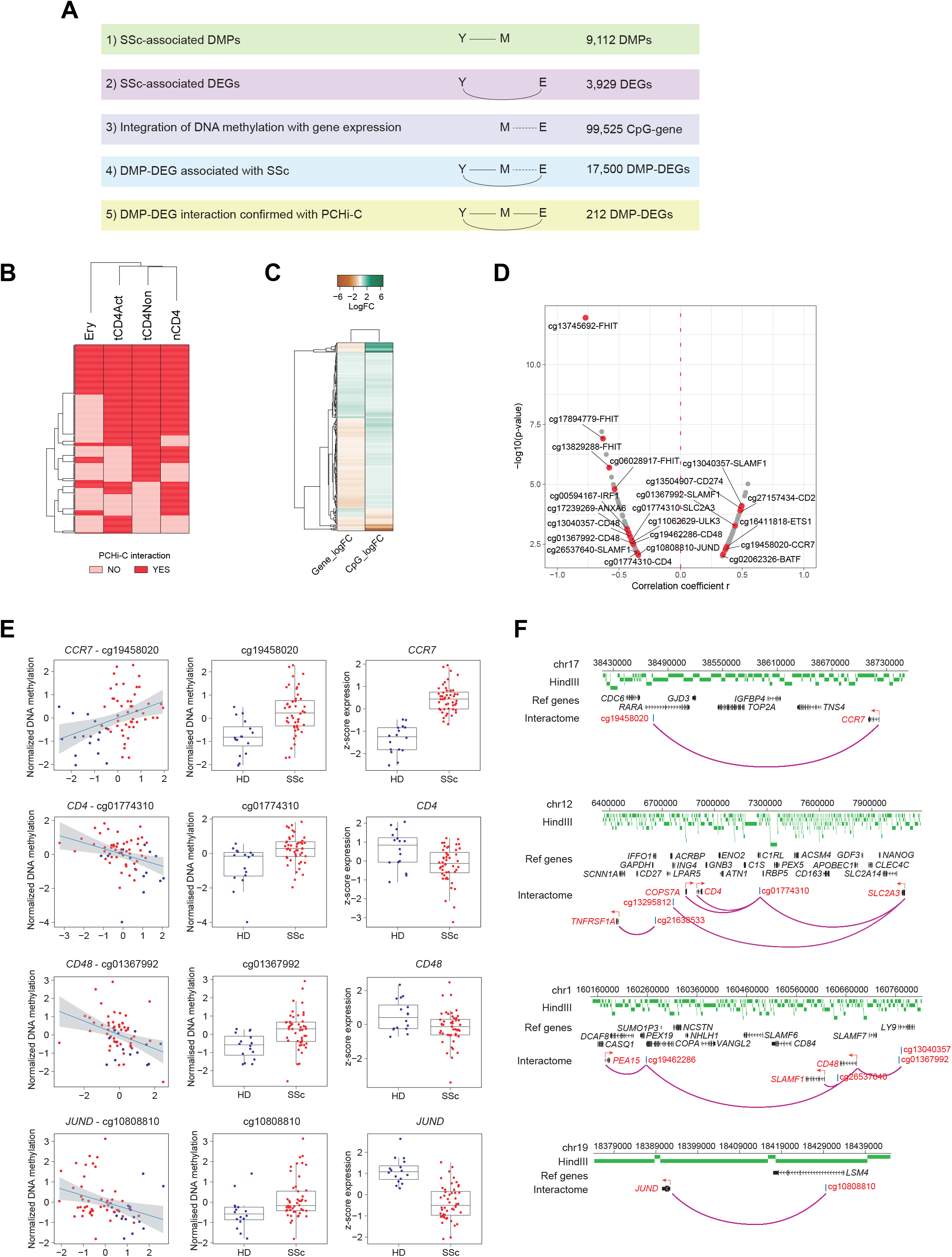
Long-range interactions as confirmed by promoter capture Hi-C (PCHi-C). (A) Correlations of possible long-range interactions between DMPs and DEGs were detected using the *MatrixEQTL* package by applying a maximum interaction distance of 5 Mb with a Pearson p-value cut-off of 0.01. Overview summarizing multi-step filtering process based on the causal inference test. Y: SSc phenotype; M: identified DMPs; E: identified DEGs; S: SSc-associated susceptibility loci. (B) Heatmap showing the presence (red) or absence (pink) of promoter capture Hi-C (PCHi-C) interactions of 212 identified DMP-DEGs identified utilizing naïve (nCD4), non-activated (tCD4Non) and activated (tCD4Act) PCHi-C datasets. The presence or absence of the same interactions were interrogated in erythroblasts (Ery). (C) Heatmap of of log_2_FC of gene expression and DNA methylation of the 212 confirmed interacting DMP-DEGs. (D) Volcano plot of the 212 confirmed DMP-DEG interactions. Correlation coefficient r of DNA methylation and gene expression values is plotted on the x-axis against –log_10_(p-value) on the y-axis. Genes relevant to T cell biology are highlighted in red. (E) Graphical representations of DMP-DEG pairs with significant correlation between DNA methylation and gene expression (Pearson p-value < 0.01). (B) Overlap between DMP-DEG pairs and PCHi-C data were performed to confirm long-range interactions. Arc plot of long distance interactions between DMPs and DEGs as confirmed by PCHi-C, in which confirmed interactions are depicted as purple arcs. HindIII fragments are depicted in green, and interacting genes and CpGs are highlighted in red.

### DMP-DEG interactions mediated by SSc-associated genetic susceptibility loci

To fully unravel the relevance of DNA methylation in SSc pathogenesis, we can hypothesize that SSc-associated genetic variants may at least partially contribute to aberrant DEG-interacting CpGs. Hence, we utilized a large GWAS dataset generated by López-Isac *et al*. ^13^, which included 26,679 individuals that identified 27 SSc-associated risk loci (S) physically interacting with 43 target genes, as validated by HiChIP data in CD4+ T cells. We therefore interrogated the presence of interacting SNPs in association with identified DMPs and DEGs. We first overlapped the SNP-interacting genomic regions with identified DEGs and observed that 36 SNP-interacting genes identified by López-Isac *et al*. were aberrantly expressed in our cohort of SSc (Figure 6A). Second, of these 36 DEGs, 5 of them interacted and correlated with a SSc-associated DMP. Finally, we observed that 4 of the identified candidate SNP-DEGs further interacted with the gene in which the associated DMP was located (Figure 6A). Specifically, the SSc-associated susceptibility loci *TNIP1* (rs3792783), *GSDMB* (rs9303277), *IL12RB1* (rs2305743) and *CSK* (rs1378942) were possible candidates that controlled both DMPs, situated in *TNIP1, RARA, LSM4* and *MPI* respectively, and their interacting DEGs, namely *ANXA6, CCR7, JUND* and *ULK3* respectively (Figure 6B). Furthermore, visualizing ChIP-seq data of CTCF and p65, obtained from the ReMap database, we observed that the SNP-DMP-DEG interaction regions were extensively covered by CTCF binding sites. Interestingly, we observed that p65 binding are predominantly restricted to the promoters/TSS of DEGs *ANXA6, CCR7* and *JUND*. Furthermore, some ChIP-seq peaks were detected near the interacting CpGs and SNPs, suggesting that p65 may play a role in the transcriptional regulation of these genes following correct chromatin looping between genomic loci (Figure 6B).

**Figure 6.**
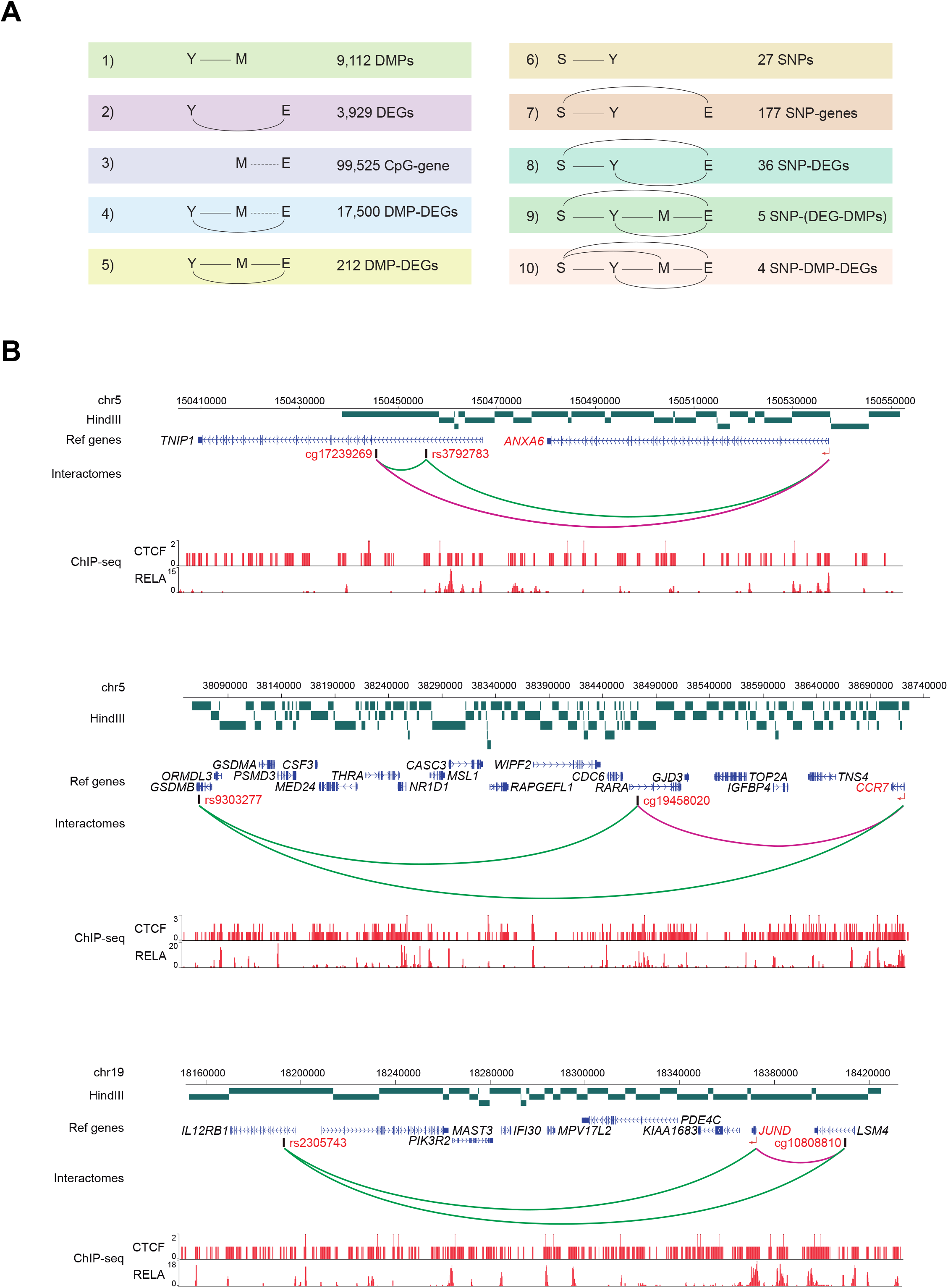
Presence of SSc-associated genetic variants interacting with PCHi-C-confirmed DMP-DEG pairs. (A) Multi-step filtering process based on the causal inference test extended from Figure 5, summarizing the filtering steps adopted to identified four interactomes involving SSc-associated SNPs, DMPs and DEGs. Y: SSc phenotype; M: identified DMPs; E: identified DEGs; S: SSc-associated susceptibility loci. (B) Arcs in green represent interactions of SSc-associated SNPs with distant genes, as confirmed by López-Isac et al. ^12^, and arcs in purple represent DMP-DEG interactions identified and confirmed in this study. HindIII fragments are depicted in green, and interacting genes and CpGs are highlighted in red. Density plots represent ChIP-seq data of RELA and CTCF. ChIP-seq data were downloaded from ReMap database, in which CTCF were downloaded as consolidated ChIP-seq peaks of all available datasets, whereas RELA ChIP-seq peaks were obtained from CD4+ T cells.

## DISCUSSION

In this study, the integrated analysis of DNA methylation, expression, promoter capture Hi-C and genetic data unveils novel functional relationships in CD4+ T cells of patients with systemic sclerosis. First, we report DNA methylation and gene expression alterations in SSc CD4+ T lymphocytes associated with essential pathways implicated in T cell differentiation and function. Aberrant DNA methylation in distinct loci could be directly influencing aberrant gene expression through long-distance interactions that involve CTCF. Finally, we identified four important SSc-associated susceptibility loci, *TNIP1* (rs3792783), *GSDMB* (rs9303277), *IL12RB1* (rs2305743) and *CSK* (rs1378942), that physically interact with cg17239269-*ANXA6*, cg19458020-*CCR7*, cg10808810-*JUND* and cg11062629-*ULK3* respectively.

Despite the increasing number of genetic variants associated with SSc, their functional relevance is still a challenge. In addition, alongside genetic predisposition, environmental factors and epigenetic deregulation also contribute to the pathogenesis of this disease (reviewed in ^48^). Our comprehensive approach has unveiled a high number of DNA methylation and expression changes in SSc CD4+ T cells compared to control (9112 DMPs, 1082 DMRs and 3929 DEGs). The vast majority of DNA methylation changes were SSc-associated hypermethylation. Conversely, only around 10 % of changes corresponded to aberrant hypomethylation, which is perhaps associated with the increased gene expression of *TET3*, as detected by our RNA expression analysis. TET3 has been previously linked to T cell differentiation and its deletion resulted in decreased proportions of progenitor and naïve CD4+ T cells ^49^. Therefore, we cannot discard the possibility that changes in DNA methylation may be a result of alterations in CD4+ T cells populations. However, deconvolution analysis showed no significant differences in the DNA methylomes of memory and naïve T cell populations between SSc and controls (results not shown).

Analysis of functional categories of the identified DMPs, DMRs and DEGs revealed the enrichment of numerous relevant pathways, in which many were in accordance to previous studies, including circadian signalling, Rho protein signalling, thyroid hormone secretion, T cell activation and cytokine-mediated responses ^50,51^, however, we were able to identify several novel pathways, including several cytokine receptor-propagated pathways, MHC complex assembly, T cell polarization and NF-κB signalling pathway, amonst others. Given the association of the HLA loci with SSc disease, as observed by previous genetic studies ^5,52^, aberrant methylation of these loci may have implications in regards to correct function of the MHC complex.

The relationship between DNA methylation and gene expression is complex, and there are studies describing DNA methylation as both a cause and a consequence of gene expression. Furthermore, DNA methylation patterns are highly tissue-specific and are established during dynamic differentiation events by site-specific remodelling at regulatory regions ^53^. Nevertheless, methylation of CpGs located in gene promoter, first exon and intron robustly correlate to gene expression in an inverse manner ^54–56^. First, we identified 45 DMPs located within or near the promoter or TSS of DEGs with statistically significant correlations, with majority displaying negative correlations. Hence, aberrant DNA methylation observed in SSc patients may directly cause aberrant gene expression in CD4+ T cells when located in/near promoter or TSS regions. Second, gene expression deregulation of several TFs were observed in SSc CD4+ T cells, including *JUN, CTCF, FLI1* and *RUNX1*, whose motifs were enriched in identified DMPs. Therefore, it is also plausible that aberrant gene expression of TFs may mediate altered recruitment to their binding sites, which may consequently shape DNA methylation patterns at these sites. Several TFs have been shown to bind unmethylated regions to block *de novo* methylation, and one such factor is CTCF, in which it acts as a boundary element to directly impede methylation of regulatory regions ^57,58^. Conversely, other TFs were observed to actively recruit DNA (de)methylation enzymes. One such example is PU.1, has been shown to recruit both TET2 and DNMT3A to promote DNA demethylation and methylation respectively ^59^.

Within the last decade, it has become increasingly clear the existence of long-range looping interactions between regulatory elements and promoters, in which only ∼7 % of all looping interactions are with the nearest gene ^60^. These interactions do not only physically exist, but play essential molecular roles in regulating distant gene expression in both biological and pathological settings ^47,61^. Long-range integration of DNA methylation and gene expression has already been explored in other disease contexts, in which one study involving a large cohort of colon cancer patients showed that methylation of distal CpGs controlled genes at a distance of > 1Mb ^62^. Furthermore, several studies have identified essential long-range associations between susceptibility loci with gene expression (GWAS) or with DNA methylation (EWAS) mediating several autoimmune diseases including multiple sclerosis ^63^, rheumatoid arthritis ^64^ and SSc ^13^. Our study represents the first to integrate genetic risk with epigenome and transcriptome deregulations, coupled with validation of physical interactions in the context of autoimmune disease. First, we identified four SSc-DMPs, cg17239269 (*TNIP1*), cg19458020 (*RARA*), cg10808810 (*LSM4*) and cg11062629 (*MPI*), whose DNA methylation correlated with the expression of four distant SSc-DEGs, *ANXA6, CCR7, JUND* and *ULK3*. Second, these four interactions were confirmed by previously published promoter-capture HiC data from CD4+ T cells ^47^. Third, from the previous study by López-Isac and colleagues, four SSc risk variants, *TNIP1* (rs3792783), *GSDMB* (rs9303277), *IL12RB1* (rs2305743) and *CSK* (rs1378942), were identified to physically interact with *ANXA6, CCR7, JUND* and *ULK3*, respectively, as well as with the genes in which the interacting SSc-DMPs, cg17239269 (*TNIP1*), cg19458020 (*RARA*), cg10808810 (*LSM4*) and cg11062629 (*MPI*), respectively, were located. Each of these identified DEGs play important roles in T cell biology. Specifically, Annexin A6 (*ANXA6*) has been described to be essential to CD4+ T cell proliferation via interleukin-2 signalling ^65^. *CCR7*, on the other hand, has been predominantly implicated in T cell migration ^66^ and its deregulation in CD4+ T cells has been associated to several autoimmune diseases including SSc ^67–69^. Finally, *JUND* encodes a TF that forms part of the AP-1 complex and has been previously described to regulate T cell proliferation and differentiation ^70^. *ULK3* encodes a serine/threonine-protein kinase that regulates SHH signalling, which is essential in T cell differentiation, proliferation and signalling ^71,72^.

One limitation of this study is we cannot accurately predict whether aberrant DNA methylation is a cause or consequence of changes in gene expression. However, given that DMPs were found to enrich in CTCF binding motifs, which may be a consequence of aberrant upregulation of the *CTCF* gene in SSc CD4+ T cells, and the importance of CTCF in enabling chromatin loop formation ^73^, it is therefore possible that aberrant DNA methylation deregulates CTCF recruitment, which has been previously described to be DNA methylation-dependent ^74^, in turn affecting long-range interactions with distant genes to alter their expression.

Collectively, our results confirm the occurrence of widespread DNA methylation and expression alterations in SSc CD4+ T cells, which at least are in part determined through long-distance interactions. Additionally, we have detected the four SSc-associated interactomes in which the presence of causal risk variants confer aberrant DNA methylation and gene expression, which consequently impacts SSc disease phenotype.

## METHODS

### Patient cohort and CD4+ T cell isolation

This study included 48 SSc patients and 16 age- and sex-matched healthy donors (HD). Individuals included in this study gave both written and oral consent in regards to the possibility that donated blood would be used for research purposes. Samples were collected at Vall d’Hebron Hospital, Barcelona, in accordance with the ethical guidelines of the 1975 Declaration of Helsinki. The Committee for Human Subjects of the Vall d’Hebron Hospital and Bellvitge Hospital approved the study. Patients with a diagnosis of SSc according to the American College of Rheumatology (ACR)/European league against rheumatism (EULAR) 2013 criteria ^75^. CD4+ T lymphocyte population was isolated from whole blood by fluorescence activated cell sorting performed at the Unitat de Biologia (Campus de Bellvitge), Centres Científics i Tecnològics, Universitat de Barcelona (Spain). Briefly, peripheral blood mononuclear cells (PBMCs) were separated by laying on Lymphocytes Isolation Solution (Rafer, Zaragoza, Spain) and centrifuged without braking. PBMCs were stained with fluorochrom-conjugated antibody against CD4-APC (BD Pharmingen, New Jersey, USA) in staining buffer (PBS with 2 mM of EDTA and 4 % FBS) for 20 mins. Gating strategies were employed to eliminate doublets, cell debris and DAPI+ cells. Lymphocytes were separated by forward and side scatter, in which CD4+ cells were separated by positive selection.

### RNA/DNA isolation

RNA and DNA were isolated from the same cell pellet utilizing AllPrep DNA/RNA/miRNA Universal Kit (Qiagen, Hilden, Germany) according to manufacturers’ instructions. RNA quality was assessed by the 2100 Bioanalyzer System (Agilent, California, USA) carried out at the High Technology Unit (UAT), at Vall d’Hebron Research Institute (VHIR).

### Illumina EPIC methylation assay and data processing

Bisulfite (BS) conversion was performed using EZ-96 DNA Methylation^™^ Kit (Zymo Research, CA, USA) according to manufacturers’ instructions. 500 ng of BS-converted DNA were hybridised on Infinium MethylationEPIC BeadChip array (Illumina, Inc., San Diego, CA, USA) following manufacturers’ instructions to assess DNA methylation of 850,000 selected CpGs that cover 99 % of annotated RefSeq genes. Fluorescence of probes was detected by BeadArray Reader (Illumina, Inc.), and image processing and data extraction were performed as previously described ^76^. Downstream data processing and normalization were performed using the R statistical language. Probes were first filtered by detection p value (p < 0.01) and normalized by Illumina normalization provided by the *minfi* package. CpGs in single nucleotide polymorphism loci were eliminated. An additional filter of beta values (ratio of DNA methylation) was applied, in which top 5 % of CpGs with the highest Δbeta between sample groups were retained for further analyses. ComBat adjustment, provided by the *sva* package, was performed to remove bias from batches. M values (log_2_-transformed beta values) were utilized to obtain p-value and adjusted p-value (Benjamini Hochberg-calculated FDR) between sample groups by an eBayes-moderated paired *t*-test using the *limma* package ^77^. P-value of < 0.01 and FDR of < 0.05 was considered statistically significant. Differentially methylated regions (DMRs) were identified using the bumphunter function from *minfi*, in which a p value of < 0.05 and a region containing 2 or more CpGs were considered DMRs.

### Clariom S gene expression array and data processing

100 ng of excellent quality RNA (RNA Integrity Number of > 9) was hybridized on Clariom^™^ S array, carried out at the High Technology Unit (UAT), at Vall d’Hebron Research Institute (VHIR), which interrogates the expression of > 20,000 transcripts. Data processing and normalization were carried out using the R statistical language. Background correction was performed using Robust Microarray Analysis (RMA) normalisation provided by *oligo* package ^78^. Annotation of probes were performed using *clariomshumantranscriptcluster*.*db* package ^79^, and the average expression level was calculated for probes mapped to the same gene. ComBat adjustment, provided by the *sva* package, was performed to remove bias from batches and other confounding variables. For comparisons between groups, the *limma* package was used to perform an eBayes-moderated paired *t*-test provided in order to obtain log_2_ fold change (log_2_FC), p-value and adjusted p-value (Benjamini Hochberg-calculated FDR). Genes that displayed statistically significant tests (p-value < 0.01 and FDR < 0.05) were considered differentially expressed.

### Gene ontology, motif and regulon enrichment analyses

Gene ontology (GO) analysis of differentially methylated CpGs (DMPs) and DMRs were performed using the GREAT online tool ((http://great.stanford.edu/public/html) ^80^, in which genomic regions were annotated by applying the basal plus extension settings. For DMPs, annotated CpGs in the EPIC array were used as background, and for DMRs, default background was used. GO terms with p-value < 0.01 and fold change (FC) > 2 were considered significantly enriched. GO analysis of differentially expressed genes (DEGs) was carried out using the online tool DAVID (https://david.ncifcrf.gov) under Functional Annotation settings, in which annotated genes in the Clariom^™^ S array were used as background. GO categories with p value of < 0.01 were considered significantly enriched.

Motif enrichment analysis of DMPs was performed using the findMotifsGenome.pl tool provided by the HOMER motif discovery software ^81^. A window of ±250 bp surrounding each DMP was applied and CpGs annotated in the EPIC array were used as background. Transcription factor (TF) enrichment of DEGs was carried out using the DoRothEA (Discriminant Regulon Expression Analysis) v2 tool ^82^. Regulons with confidence score of A-C were utilized for analysis and a p value of < 0.05 and Normalized Enrichment Score (NES) of ± 2 was considered significantly enriched.

### Comparative analysis with public ChIP-seq datasets

DNase I hypersensitivity and ChIP-seq data of histone modifications H3K27ac, H3K4me1, H3K4me3, H3K27me3, H3K36me3 and H3K9me3 of total CD4+ T cells were downloaded from the BLUEPRINT portal (http://dcc.blueprint-epigenome.eu/). Five independent datasets were downloaded for each histone mark and ChIP-seq peaks were consolidated using the MSPC program ^83^, in which peaks were first filtered by q.value < 0.01 and FC >= 2 and using the parameters -w = 1E^-4^, -s =1E^-8^, and –c = 3.

ChIP-seq datasets of transcription factors were downloaded from ReMap database (http://pedagogix-tagc.univ-mrs.fr/remap). ChIP-seq data of STAT1, RUNX1, NFKB2, MYC, JUN, CTCFL and CTCF were downloaded as merged peaks of all available data, whereas RELA data was generated in CD4+ T cells. Background of DMRs were generated from annotated CpGs from EPIC array by randomly permutating 1000 times, and an average p value of < 0.01 and average odds ratio > 1 were considered significantly enriched.

### Methylation-expression quantitative trait loci (meQTL) and promoter capture Hi-C (PCHi-C) data analyses

Methylation-expression quantitative trait loci (meQTL) analysis to link varying gene expression to aberrant DNA methylation was carried out utilizing the *MatrixEQTL* package, including sex and age as covariates ^84^. A window of 5 Mb was applied for *cis* interactions and a Pearson correlation p-value of < 0.01 was considered significant. CpG-gene interactions were then filtered by differential methylation and expression in SSc T cells compared to controls (FDR < 0.05 and p-value < 0.01). Interactions were further confirmed by overlapping with PCHi-C datasets generated in naïve, non-activated and activated CD4+ T cells ^47^. Utilizing significant interactions, we first performed overlap between DEGs and mapped promoters from PCHi-C datasets. Second, we utilized pipelines provided by *GenomicRanges* package ^85^ to overlap the interacting PCHi-C fragments with DMP coordinates. Finally, we eliminated the DMPs found in gene promoters, in which 212 unique DMP-DEG interactions remained. Confirmed interactions were visualized using WashU Epigenome Browser (http://epigenomegateway.wustl.edu/legacy).

### Association analysis of interacting DMP-DEGs with GWAS data

Genome-wide association study (GWAS) data was obtained from López-Isac *et al*. ^13^, in which SSc-associated susceptibility loci and their interacting genes were overlapped, by gene name, with DMP-DEG pairs identified from this study.

## Data Availability

GEO Series accession number GSE146093.

## DATA ACCESS

DNA methylation and expression data for this publication have been deposited in the NCBI Gene Expression Omnibus and are accessible through GEO Series accession number GSE146093.

## Author contributions

T.L., E.B. and J.M. conceived and designed the study; T.L. and L.C. performed sample preparation and purification; T.L., L.O. E.A.-L., B.M.J. E.L.-I. performed bioinformatic analyses; A.G.C. and C.S.-A. provided patient samples and analyzed clinical data; E.B. and J.M. supervised the study; T.L., E.B. and J.M. wrote the manuscript; all authors participated in discussions and interpreting the results.

